# Development and Validation of Suspicious Liver Lesion Deep Learning Algorithm for Computed Tomography

**DOI:** 10.1101/2021.12.15.21267873

**Authors:** Jacob Johnson, Kaneel Senevirathne, Lawrence Ngo

## Abstract

In this work, we report the results of a deep-learning based liver lesion detection algorithm. While several liver lesion segmentation and classification algorithms have been developed, none of the previous work has focused on detecting suspicious liver lesions. Furthermore, their generalizability remains a pitfall due to their small sample size and sample homogeneity. Here, we developed and validated a highly generalizable deep-learning algorithm for detection of suspicious liver lesions. The algorithm was trained and tested on a diverse dataset containing CT exams from over 2,000 hospital sites in the United States. Our final model achieved an AUROC of 0.84 with a specificity of 0.99 while maintaining a sensitivity of 0.33.

## Introduction

Incidental liver lesions are findings identified in patient imaging that are otherwise not causing any relevant symptoms. Although most lesions are benign, a significant subset represents yet undiagnosed malignancy. In an oncologic population, about 12% of small incidental lesions represent metastatic disease (1). In non-oncological populations, other risk factors, such as chronic liver disease, can significantly increase the chances that such lesions represent primary hepatic malignancies (2). Thus, diagnosis and characterization of the type and the nature of the lesion is crucial as treatment and management approaches greatly vary.

Computed tomography (CT) medical imaging exams are a common method for identifying incidental liver lesions. According to previous reports, liver lesions can be seen in up to 12-13% of CT exams (1). However, despite close examination by radiologists, several technical and non-technical factors could complicate the detection of such incidental lesions and could be overlooked.

Contrast phase is significant in determining whether a lesion can be detected. Different hepatic lesions require different and often multiple phases of contrast for detection and/or characterization of incidental liver lesions. Since most CT examinations are performed as either non-contrast or single-phase contrast exams, this can make the task of detection or characterization particularly difficult. The relative similarity of attenuation of many hepatic lesions to the surrounding hepatic parenchyma further complicates detection. One strategy that may help with detection of these lesions is using tighter window-level settings (liver window) for visualization (3). However, the use of such settings can be variable, especially if the abdomen is not the primary anatomy of interest for a scan (e.g., chest CTPA).

Furthermore, providing rapid and accurate interpretations of imaging can be challenging for radiologists due to the increased work hours and study volumes (4,5). Studies have shown that this increases the error rates in radiology (6,7). Therefore, human error may also lead to missed incidental liver lesions. Quality assurance programs may be one approach in improving the detection of incidental hepatic lesions across radiology practices (8). However, such programs can be ineffective given the overall low rate of discrepancies found in peer review (9). There is growing literature on the use of AI in the detection and segmentation of liver lesions (10–13), and we aim to extend these efforts by building an algorithm with the eventual goal of identifying missed liver lesions.

## Methods

We performed a retrospective analysis of CT examinations, including some portions of the liver spanning from March 2017 to December 2020 from Virtual Radiologic’s (vRad) practice. All the CT imaging and radiologic data were fully anonymized in a Health Insurance Portability and Accountability Act (HIPAA) compliant manner using proprietary software from vRad. This study was approved with a waiver of consent by an external Institutional Review Board, Western IRB.

A total of 38,273 bounding box annotations and 45,514 background images (images without suspicious liver lesions) from CT scans were used for training and validation (with a 5:1 train-validation split). Bounding box annotations and segmentations of liver lesions for training and validation were made by radiologists with at least 5 years of radiology experience. For the purposes of this study, a suspicious liver lesion would be any lesion with an average Hounsfield unit attenuation of greater than 20, size greater than 1.5 cm, and no other imaging feature typical of a benign lesion (such as peripheral nodular enhancement consistent with hemangioma).

We developed a proprietary ensemble artificial intelligence visual classifier (hereafter called VC) to detect suspicious liver lesions. The VC ensemble model included (1) an initial object detector built on the YOLOv5 architecture (14), (2) a liver segmentation algorithm based on the U-Net architecture (15), and (3) a liver lesion segmentation algorithm also based on the U-Net architecture. The liver segmentation model was trained on 4,127 images from 268 unique studies with liver masks that were manually labeled by a radiologist to separate the organ from the background. Filtering by HU and lesion size was done by a lesion segmentation model. After initial object detection, potential lesion patches were extracted from the image and segmented by a separate model based on the U-Net architecture. This model was trained on 994 images from 71 unique studies that were manually labeled by a radiologist to separate the lesion from the background. After segmentation of the lesion the mean HU value is determined. Subsequently, an ellipse is fit to the segmentation and the lesion dimension is determined as the major axis of the fitted ellipse. On a per-lesion basis, a threshold was drawn at >20 HU and 1.5 cm for attenuation and size respectively. Any lesions detected by the earlier stage of the ensemble which did not meet these thresholds were classified as negative for a suspicious liver lesion. The maximum confidence score from a YOLOv5 bounding box detection was taken as the patient level confidence score for a given patient. In sum, the VC ensemble model inputs a set of Digital Imaging and Communications in Medicine (DICOM) files and generates a patient level confidence score of whether an exam is positive or negative for liver lesions.

Additionally, a unique set of 3,568 CT examinations was used as the test set. These studies were consecutive studies from a subset of exams from June 2017 at Virtual Radiologic’s practice. Data in this set was obtained from a larger distribution spanning across the United States from more than 2,000 hospital sites. The dataset also represented a variety of commercially available brands and models of CT scanners. The test data set was 57% female with patients aged 12 to 99 years (mean 57.1). This was done by first labeling whether or not reports mentioned a suspicious liver lesion as defined by our criteria (Supplemental). Reports that did not mention a suspicious liver lesion but were flagged as being positive by a preliminary version of our visual classifier were reviewed by a radiologist. Studies that were initially negative by report and positive by secondary radiologist review were given a positive ground-truth label. Of the test CT examinations, 117 cases were identified as suspicious for liver lesions.

Finally, we evaluated the performance of the VC ensemble model on the test set by measuring the sensitivity, specificity, positive predictive value (PPV), and the negative predictive value (NPV). We also measured the receiver operating characteristic (ROC) curve and the area under the ROC curve (AUROC) to assess the model further.

## Results

The incidence of liver lesions within the test set was 0.03 (117 positive and 3,451 negative). The VC ensemble model AUROC was 0.84. To identify liver lesions with high confidence, we used a high specificity threshold of 0.69. Using this threshold, we achieved a specificity of 0.99 and positive predictive value of 0.53. The sensitivity of the VC ensemble model was 0.33. The summary of our model performance for the detection of incidental liver lesions on the test set is shown in Table 1 and Figure 1.

**Table 1:** Performance summary of the Visual Classifier Ensemble Model.

|  | Incidental Liver Lesion |
| --- | --- |
| AUROC | 0.84 |
| Sensitivity | 0.33 |
| Specificity | 0.99 |
| PPV | 0.53 |
| NPV | 0.98 |

**Figure 1:**
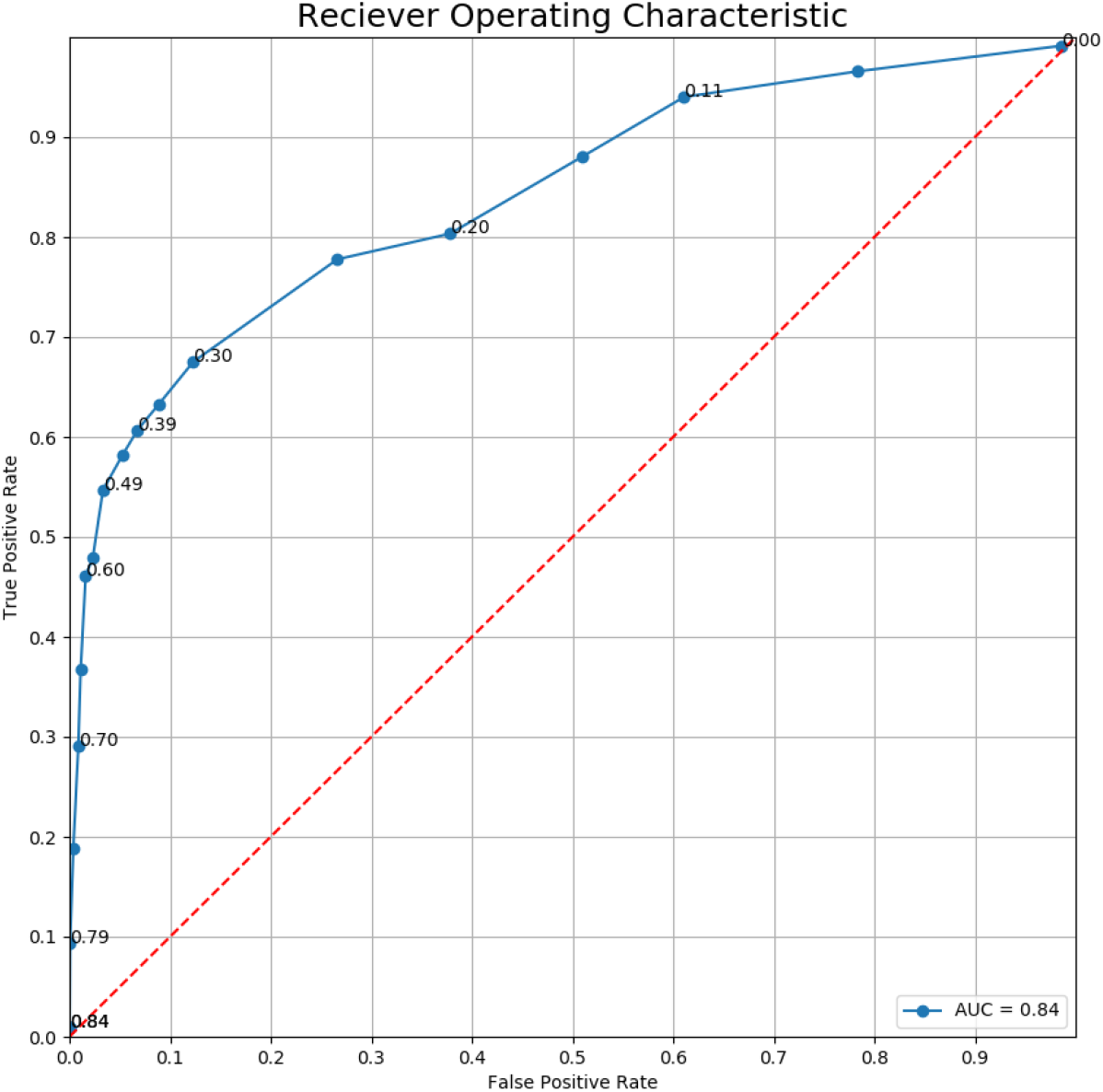
Performance of the Visual Classifier Ensemble Model for classification for presence or absence of liver lesions

## Discussion

In this study, we have implemented a deep learning based visual classifier model to detect suspicious liver lesions using CT exams of patients. We have used a high decision confidence threshold to increase the specificity of our model thereby maximizing the usability in a clinical setting. We examined this aim by testing our model in a vastly distributed test data set and achieving a specificity of 0.99. The higher specificity reduces the falsely identified positive cases. Furthermore, our test data set was obtained using a wide variety of commercially available scanners which makes our VC ensemble classifier applicable in different clinical settings without any constraints.

Several previous studies have used deep neural networks for classification and segmentation of liver lesions (10–13). Most of these models are based on deep convolutional neural networks and trained on small and homogeneous datasets. To overcome the small sample size, some of these studies have used synthetic data augmentation and generation techniques, such as Generative Adversarial Networks (GANs) (13). Although these studies have reported high sensitivity and specificity values, clinical usage of these models is questionable due to the heterogeneity of data samples. In our study we used a much bigger and diverse dataset. For instance, test CT exams were generated by multiple commercially available scanners from several vendors, such as GE (1209), Siemens (656), Philips(293), Toshiba (363) and Hitachi (4). Therefore, our model will likely be generalizable and usable in a clinical setting.

Furthermore, in previous studies, several model architectures have been implemented for classification, detection and segmentation of liver lesions. However, our VC ensemble model has multi-stage architecture that includes both an object detection phase built on the YOLOv5 architecture and a segmentation phase built on the U-Net architecture prior to classification. Thus our model is able to stratify lesions based on their level of suspicion for malignancy based on size and attenuation.

In the current study, the diagnosis (suspicious for liver lesion or not) is taken from a report generated by radiologists. However, there is the possibility that radiologists misclassify images. Despite our efforts to alleviate this by manually adjusting misclassified CT exams (supplemental), errors still could persist in our dataset. Thus, this may have an influence on the learning of the VC ensemble model and the metrics for reported test set accuracy. Thus, additional work is aimed towards reviewing VC findings against radiologists errors. In order to avoid the infeasibility of going over thousands of exams manually, we have also developed a NLP algorithm with sufficient accuracy to detect discrepancies between the VC ensemble model findings and reports generated by radiologists. We aim to apply the combination of algorithms for the purposes of finding diagnostic errors in the detection of suspicious liver lesions in future work.

## Data Availability

The data from this manuscript is proprietary and not publicly available.

## Acknowledgements

We would like to thank Nadine Ly for her assistance in paper draft editing, Aleen Altamirano for providing a subset of radiology annotations, and Brian Baker, Christine Lamoureux, Ian Driscoll, and Jerry Lohr from Virtual Radiologic for their support with clinical data.

## Supplemental Methods

NLP criteria for a suspicious liver lesion:

- Positive if the report mentions:
  - Imaging is recommended unequivocally to further evaluate a liver lesion
  - Report mentions a cystic mass.
  - Liver lesion that is indeterminate with or without follow up recommendation.
  - Suspected hemangioma but recommend follow up
  - Something in findings related to a suspicious liver lesion but never recommends anything for it. Report does not call it indeterminate.
  - Known suspicious liver lesions that may or may not be described in more detail.
- Negative if the report mentions:
  - Cyst, too small to characterize.
  - Lesion favored to be a cyst.
  - Stable hypodensity with no other description.
  - Small hypodensity not otherwise characterized.
  - Liver granulomas.
  - Lipoma.

